# Neutralizing immunity in vaccine breakthrough infections from the SARS-CoV-2 Omicron and Delta variants

**DOI:** 10.1101/2022.01.25.22269794

**Authors:** Venice Servellita, Abdullah M. Syed, Noah Brazer, Prachi Saldhi, Miguel Garcia-Knight, Bharath Sreekumar, Mir M. Khalid, Alison Ciling, Pei-Yi Chen, G. Renuka Kumar, Amelia S. Gliwa, Jenny Nguyen, Alicia Sotomayor-Gonzalez, Yueyuan Zhang, Edwin Frias, John Prostko, John Hackett, Raul Andino, Jennifer Doudna, Melanie Ott, Charles Y. Chiu

**Author notes:** Equal contribution (Venice Servellita, Abdullah M. Syed, Noah Brazer, Prachi Saldhi).

## Abstract

Virus-like particle (VLP) and live virus assays were used to investigate neutralizing immunity to Delta and Omicron SARS-CoV-2 variants in 239 samples from 125 fully vaccinated individuals. In uninfected, non-boosted individuals, VLP neutralization titers to Delta and Omicron were reduced 2.7-fold and 15.4-fold, respectively, compared to wild-type (WT), while boosted individuals (n=23) had 18-fold increased titers. Delta breakthrough infections (n=39) had 57-fold and 3.1-fold titers whereas Omicron breakthrough infections (n=14) had 5.8-fold and 0.32-fold titers compared to uninfected non-boosted and boosted individuals, respectively. The difference in titers (p=0.049) was related to a higher proportion of moderate to severe infections in the Delta cohort (p=0.014). Correlation of neutralizing and spike quantitative antibody titers was decreased with Delta or Omicron compared to WT. Neutralizing antibodies in Delta and Omicron breakthrough infections increase overall, but the relative magnitude of increase is greater in more clinically severe infection and against the specific infecting variant.

## Introduction

Variants of concern have emerged throughout the COVID-19 pandemic, causing multiple waves of infection (Dyson *et al*. 2021). The Omicron (B.1.1.529) variant has been shown to be highly transmissible with decreased susceptibility to therapeutic monoclonal antibodies and neutralizing antibodies conferred by vaccination or prior infection (Flemming 2022; VanBlargan *et al*. 2022). These characteristics are likely due to a large number of mutations in the spike protein (n=30) (Cao *et al*. 2021). Omicron has spread to become the predominant circulating lineage worldwide as of early January 2022 amidst high background levels of Delta (B.1.617.2) variant infection (Gangavarapu *et al*., 2020). The surge in Omicron has led to a reinstatement of public health interventions to decrease transmission and a renewed focus on vaccination efforts, although evidence to date suggests that Omicron causes less severe disease than other SARS-CoV-2 variants (Wolter *et al*. 2022; Davies *et al*. 2022).

The development of neutralizing antibody responses in Delta and Omicron breakthrough infections remain largely unexplored. Here we evaluated neutralizing antibody responses in fully vaccinated individuals, some of whom were boosted and/or subsequently developed a COVID-19 breakthrough infection. Neutralization was assessed using two independent methods that incorporated either SARS-CoV-2 virus-like particles (VLP) incorporating all of the Omicron mutations in the spike, nucleocapsid, matrix, and fusion structural proteins (Syed *et al*. 2021; Syed *et al*. 2022) or live viruses in the assay. We also correlated the neutralization results with quantitative spike antibody levels and investigated associations between neutralizing antibody titers and infecting variant or clinical severity of the breakthrough infection.

## Results

### Neutralizing antibody levels in vaccinated individuals wane over time and are reduced against the Delta and Omicron variants

VLP and live virus neutralization assays were performed in parallel on 144 plasma samples collected from 81 subjects enrolled in a prospectively enrolled longitudinal cohort (the UMPIRE, or “UCSF eMPloyee and community Immune REsponse study”), 18 (22.2%) of whom had received a booster and none of whom were previously infected. We chose available samples from the earliest and most recent time points collected from each subject ≥14 days after the last vaccine dose for neutralization testing. Sample collection dates for fully vaccinated, non-boosted individuals (n=63) ranged from 14 to 305 days (median = 91 days) following completion of the primary series of 2 doses for an mRNA vaccine (BNT162b2 from Pfizer or mRNA-1273 from Moderna) or 1 dose of the adenovirus vector vaccine (Ad26.COV2.S from Johnson and Johnson); for boosted individuals (n=18), collection dates ranged from 2 to 74 days (median = 23 days) following the booster dose. Overall, median neutralizing antibody titers were 2.5-fold lower using live viruses compared to VLPs **(Supplementary Figure 1)**.

In non-boosted vaccinated individuals, median VLP neutralizing antibody titers to Delta and Omicron relative to the ancestral wild-type (WT) virus WA-1 lineage were reduced 2.7-fold (262 ⟶ 17,expressed as NT50 titers, or titers needed to neutralize 50% of VLP activity) and 15.4-fold (262 ⟶ 96), respectively **(Figure 1A and B)**. In comparison, live virus neutralization titers to Delta and Omicron were both reduced 3.0-fold (120 ⟶ 40), with the lower reduction for Omicron accounted for by the higher limit of detection (LOD) for the live virus (NT50 = 40) compared to the VLP neutralization assay (NT50 = 10). Using VLPs, the proportion of individuals with neutralizing antibodies to Omicron above an NT50 cutoff of 40 was ∼20%, as compared to ∼80% and ∼95% for Delta and WT, respectively **(Figure 1C, left)**. The corresponding proportions using live viruses were ∼5%, ∼45%, and ∼75% for Omicron, Delta, and WT, respectively **(Figure 1C, right)**. In boosted individuals, baseline VLP titers to WT were 18-fold higher (4,727 versus 262) than in non-boosted individuals **(Figure 1A, B, D, and E, left)**, and decreases in titers to Delta and Omicron relative to WT were more modest at 3.3-fold and 7.4-fold, respectively **(Figure 1D and E, left)**. The increase in VLP neutralization titers corresponded to >93% of boosted individuals having neutralizing antibodies to all 3 lineages above an NT50 cutoff of 40 **(Figure 1F, left)**. In contrast, live virus neutralization titers in boosted individuals showed a 21.4-fold (1,475⟶69) decrease in titers to Omicron relative to WT **(Figure 1E, right)**, with only ∼62% of boosted individuals having neutralizing antibodies to Omicron **(Figure 1F, right)**. Median VLP neutralization titers to WT decreased by 93% (14-fold, 2,043⟶146) over >3 months following vaccination, with decreases in titers to Delta and Omicron of 2.9-–4.7-fold and 12.2-43.5-fold, respectively, relative to WT **(Figure 1G)**.

**Figure 1.**
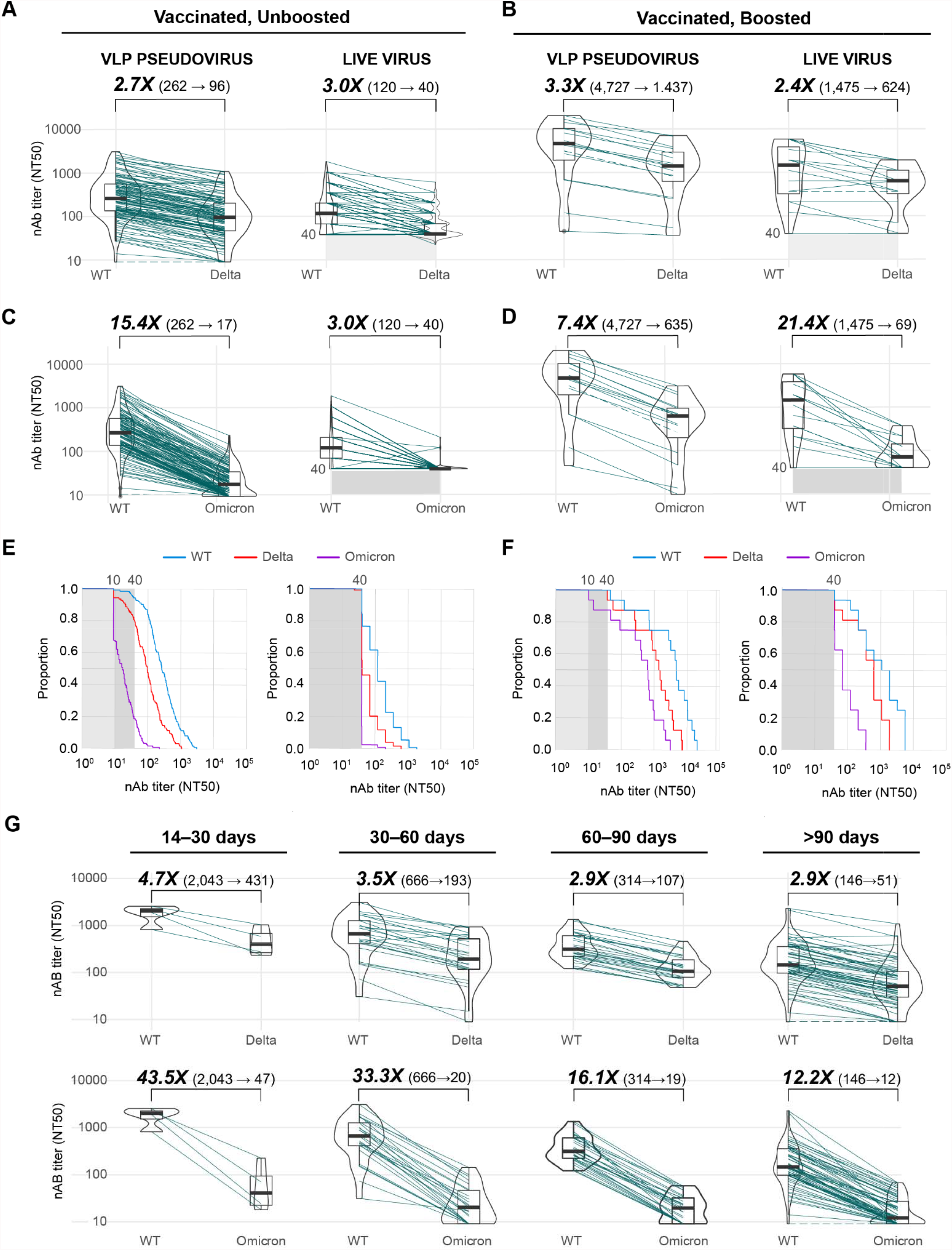
Neutralizing antibody levels in fully vaccinated, uninfected individuals. **(A, B)** Box-violin plots showing median neutralizing antibody titers using VLP (left) and live virus (right) assays against the SARS-CoV-2 WA-1 ancestral lineage (wild-type, or “WT”) and Delta variant in vaccinated, unboosted **(A)** and vaccinated, boosted **(B)** individuals **(C, D)** Box-violin plots of titers against the WT and Omicron variant in vaccinated, unboosted **(C**) and vaccinated, boosted **(D)** individuals. **(E, F)** Cumulative distribution function plots of titers to WT, Delta, and Omicron using VLP (left) and live virus (right) assays in vaccinated, unboosted **(E)** and vaccinated, boosted **(F)** individuals, showing the proportion of samples at or above a given titer. **(G)** Longitudinal box-violin plots of VLP titers to Delta (top) and Omicron (bottom) stratified by time ranges following completion of a primary vaccine series.

### Breakthrough infection increases baseline neutralizing antibody levels and variant-specific immunity

To investigate neutralizing antibody responses and the extent of cross-variant immunity, we analyzed plasma samples from 53 patients with confirmed SARS-CoV-2 breakthrough infections. Of the 53 cases, 28 and 14 were identified as Delta and Omicron variants, respectively, by viral whole-genome sequencing. For the remaining 11, we were unable to identify the lineage because of lack of a respiratory swab sample or insufficient genome coverage for definitive identification. These 11 were presumptively identified as Delta because they were collected from July 30 to December 1, 2021, during a period of when Delta accounted for 97.1 – 99.6% of the circulating lineages in California (CDPH 2022). The number of days between sample collection and symptom onset or PCR positivity, whichever was earlier, ranged from 1 to 55 days (median = 15 days). Of the 53 breakthrough cases, 28 (52.8%) were hospitalized with moderate to severe COVID-19 disease and 13 (24.5%) were immunocompromised.

Using VLP assays, we found that Delta breakthrough infections (n=39), 5 of which were boosted, had increased median baseline WT neutralization titers of 57-fold (14,835 versus 262) and 3.1-fold (14,835 versus 4,727) compared to those from non-boosted and boosted individuals, respectively **(Figure 1A, top and 2A, top)**. In addition, neutralization titers to Delta rose to the same level as WT in the live virus assay **(Figure 2A, bottom)**. Partial neutralization to Omicron was observed mainly due to the increase in baseline titers, as there appeared to be limited cross-variant immunity with 31.1-fold and 46.8-fold decreases in Omicron neutralization relative to WT for the VLP and live assays, respectively **(Figure 2A)**. The proportion of individuals with neutralizing antibodies to Omicron above an NT50 cutoff of 40 was calculated at ∼75% **(Figure 2A, top right)** and ∼43% **(Figure 2A, bottom right)** for the VLP and live virus assays, respectively.

**Figure 2.**
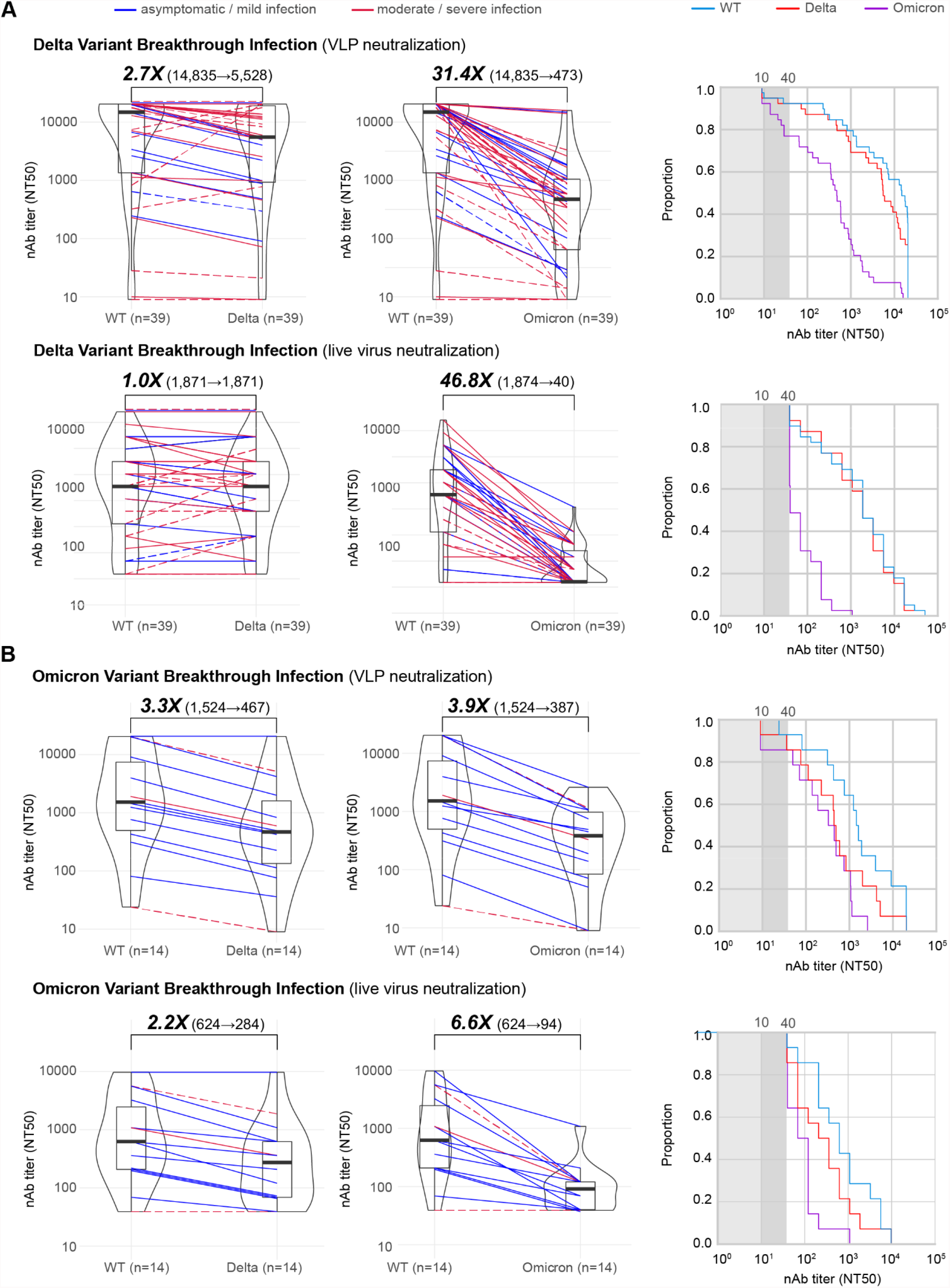
Neutralizing antibody levels in Delta and Omicron breakthrough infections. **(A)** Box-violin plots of median neutralizing antibody titers against Delta (left) and Omicron (middle) variants compared to WT, along with cumulative distribution function plots of titers to WT, Delta, and Omicron (right), showing the proportion of samples at or above a given titer, in patients with Delta breakthrough infections using VLP (top row) and live virus (bottom row) assays. **(B)** Corresponding box-violin and cumulative distribution plots in patients with Omicron breakthrough infections. For the box-violin plots, the median is represented by the thick black line inside the box. The lines connecting the paired points are color-coded based on severity of infection (blue = asymptomatic or mild infection, red = moderate or severe infection). The solid lines denote immunocompetent and the dashed lines immunocompromised patients.

In contrast to Delta, Omicron breakthrough infections (n=14), of which only 1 was boosted, exhibited much smaller increases in baseline WT titers, 5.8-fold those from non-boosted individuals and only to about one-third of the titers from boosted individuals **(Figure 1A and 2B, top left)**. However, a relative increase in neutralization titers to Omicron was observed, with Omicron-specific titers in breakthrough infections only 3.9-fold lower than WT **(Figure 2B, top left)**. In contrast, no corresponding increase in Delta-specific titers was observed, with a 3.3-fold reduction in titer that was comparable to that observed previously in uninfected vaccinated individuals **(Figure 1A and 2B, top left)**. Thus, Omicron breakthrough infection resulted in ∼90% of individuals having neutralizing antibodies to Omicron above an NT50 cutoff of 40, comparable in proportion to those having neutralizing antibodies to Delta (**Figure 2B, bottom left)**.

A head-to-head comparison of Omicron and Delta breakthrough infections was performed after inclusion of samples collected from 0 to 30 days after symptom onset **(Figure 2B, left and right)**. The comparison showed that Delta breakthrough infections resulted in more pronounced increases in baseline WT titers relative to non-boosted vaccinated individuals, 61.8-fold (16,195 versus 262) versus 5.8-fold (1,524 versus 262) **(Figure 1A and 2B)**. The difference in median neutralization titers between Delta and Omicron variants was significant when including all breakthrough patients (9.7-fold, 14,835⟶1,524, p=0.049) and when considering only immunocompetent patients (13.1-fold, 19,921⟶1,524, p=0.015) **(Figure 3A and 3B, left)**. While the breakthrough infection conferred additional neutralizing immunity against the infecting variant **(Figure 1A, bottom and Figure 1B, left)**, the extent of cross-variant immunity beyond increases in baseline WT titers was limited, with a 3.3-fold reduction in Delta titers in Omicron breakthroughs and a 36.1-fold reduction in Omicron titers in Delta breakthroughs **(Figure 2A and B)**, comparable to the reductions observed in uninfected vaccinated individuals (**Figure 1A)**.

**Figure 3.**
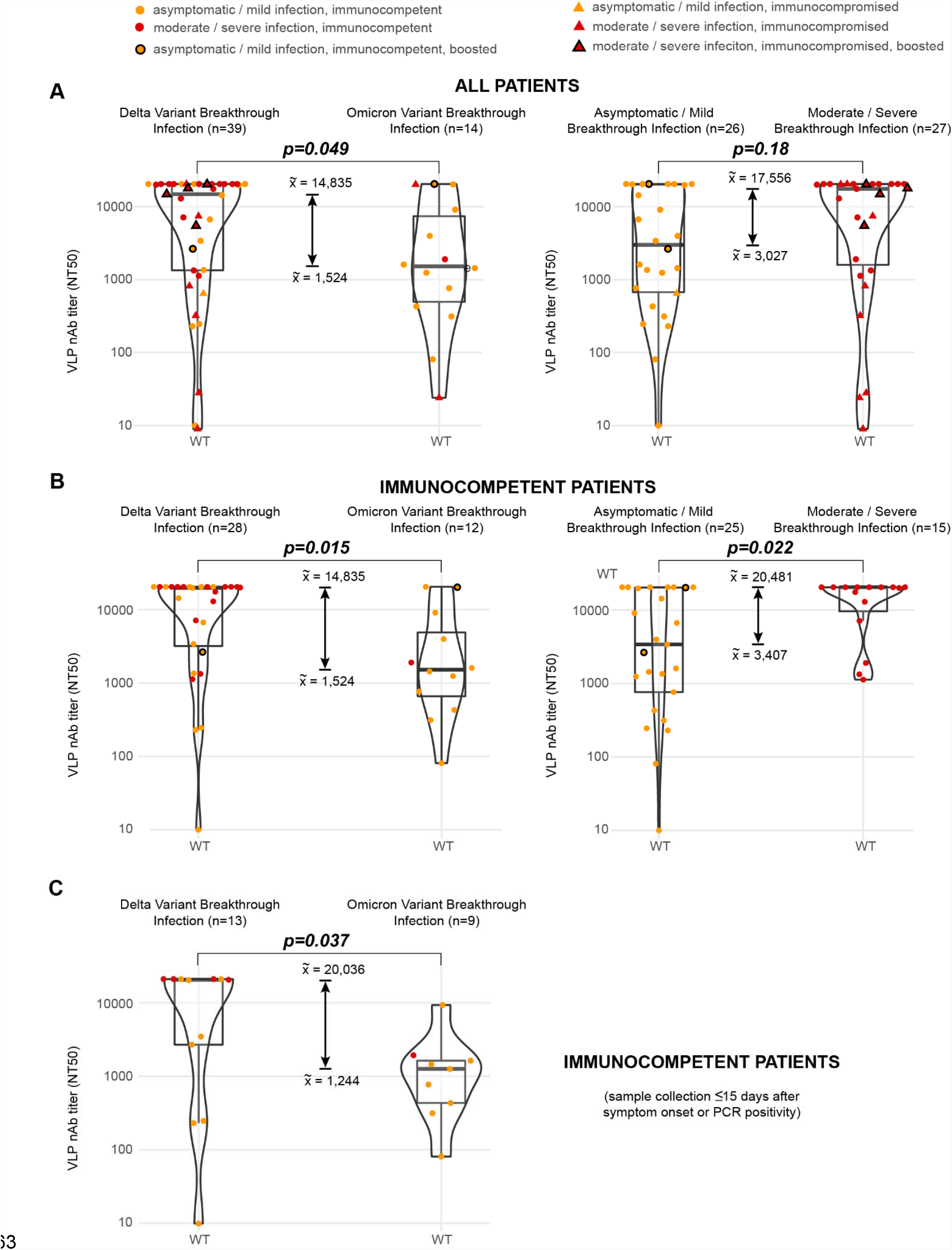
Comparison of neutralizing antibody titers against the WT lineage in Delta and Omicron breakthrough infections. **(A)** Box-violin plots comparing neutralizing antibody titers against the WT lineage between Delta and Omicron breakthrough infections (left) and between asymptomatic or mild and moderate or severe breakthrough infections (right) for all patients in the study. **(B)** Corresponding box-violin plots for immunocompetent patients only. **(C)** Box-violin plots between Delta and Omicron breakthrough infections for samples that were collected ≤15 days after symptom onset or PCR positivity. P-values for significance were determined using the Mann-Whitney U test.

### Clinical severity is a potential determinant of neutralizing immunity

We sought to identify clinical factors that may explain the difference between neutralizing antibody levels between Delta and Omicron breakthrough infections. Univariate analysis revealed that among the factors examined, only clinical severity, hospitalization for COVID-19, and median numbers of days between symptom onset or PCR positivity and sample collection, were significant **(Table 1)**. Notably, there was a higher proportion of moderate to severe infections in Delta compared to Omicron breakthrough cases **(Table 1, p=0**.**014)**. In immunocompetent patients, moderate to severe breakthrough infections from Delta and Omicron were found to elicit higher median levels of neutralizing antibodies as compared to mild or asymptomatic infections **(Figure 3B, right, 20**,**481 versus 3**,**407, p=0**.**022)**. The corresponding comparison was not significant for all patients **(Figure 3A, right, 17**,**556 versus 3**,**027, p=0**.**18)**, as there were several vaccinated patients in the current study who were hospitalized for moderate to severe COVID-19 breakthrough infection yet failed to generate a robust antibody response due to their immunocompromised state **(Figure 3A, red triangles)**. Since the median number of days between symptom onset or PCR positivity and sample collection for Delta and Omicron infections was also significant **(Table 1, 17 versus 8**.**5 days, p=0**.**008)**, we performed a sub-analysis of neutralization titers between Delta and Omicron breakthrough infections for samples collected ≤15 days from symptom onset or PCR positivity. The difference in levels of neutralizing antibodies from Delta or Omicron breakthrough infection was still significant **(Figure 3C, right, p=0**.**037)**, even at comparable time intervals between symptom onset or PCR positivity and sample collection **(Table 1, 9 versus 6 days, p=0**.**12)**.

**Table 1.** Clinical and demographic characteristics in Delta and Omicron variant breakthrough infections. P-values for significance were determined using two-tailed Fisher’s Exact Test for the categorical variables and the Mann-Whitney U test for the days between sample collection and symptom onset or PCR positivity.

| Characteristic |  | Delta<br>variant | Delta<br>variant(%) | Omicron<br>variant | Omicron<br>variant(%) | p-value | lower<br>95%CI | upper<br>95%CI | odds ratio |
| --- | --- | --- | --- | --- | --- | --- | --- | --- | --- |
| gender | female | 17 | 43.59% | 8 | 57.14% | 0.5343 | 0.14 | 2.35 | 0.59 |
|  | male | 22 | 56.41% | 6 | 42.86% |  |  |  |  |
| age | >65 | 19 | 48.72% | 6 | 42.86% | 0.7632 | 0.31 | 5.32 | 1.26 |
|  | 18-65 | 20 | 51.28% | 8 | 57.14% |  |  |  |  |
| received booster | yes | 5 | 12.82% | 1 | 7.14% | 1 | 0.18 | 97.23 | 1.89 |
|  | no | 34 | 87.18% | 13 | 92.86% |  |  |  |  |
| severity | moderate to severe | 24 | 61.54% | 3 | 21.43% | 0.01351 | 1.23 | 36.82 | 5.67 |
|  | asymptomatic to mild | 15 | 38.46% | 11 | 78.57% |  |  |  |  |
| hospitalized for covid | yes | 25 | 64.10% | 3 | 21.43% | 0.01116 | 1.36 | 41.17 | 6.31 |
|  | no | 14 | 35.90% | 11 | 78.57% |  |  |  |  |
| immune status | immunocompromised | 11 | 28.21% | 2 | 14.29% | 0.4729 | 0.41 | 24.74 | 2.32 |
|  | immunocompetent | 28 | 71.79% | 12 | 85.71% |  |  |  |  |
| median collection interval from symptom onset |  |  |  |  |  |  |  |  |  |
| all samples |  | 17 days |  | 8.5 days |  | 0.008 |  |  |  |
| ≤ 15 days |  | 9 days |  | 6 days |  | 0.1231 |  |  |  |

### Quantitative spike antibody assays show decreased correlation with and are less predictive of neutralizing activity against the Delta and Omicron variants

We compared VLP and live virus neutralization with quantitative spike antibody results from an FDA Emergency Use Authorization (EUA) authorized commercial assay **(Figure 4)**. The results showed decreased correlation of neutralization and quantitative antibody titers with Omicron (Spearman’s ρ=0.49-0.75) and Delta (ρ=0.83-0.88) relative to WT (ρ=0.91-0.93). Of note, many cases of Delta breakthrough infection with low to moderate levels of spike IgG antibody failed to neutralize Omicron in the live virus assay **(Figure 4B, bottom)**. Quantitative NT50 titers of 10^3^–10^4^ and >10^5^ reliably predicted Delta and Omicron neutralization, respectively.

**Figure 4.**
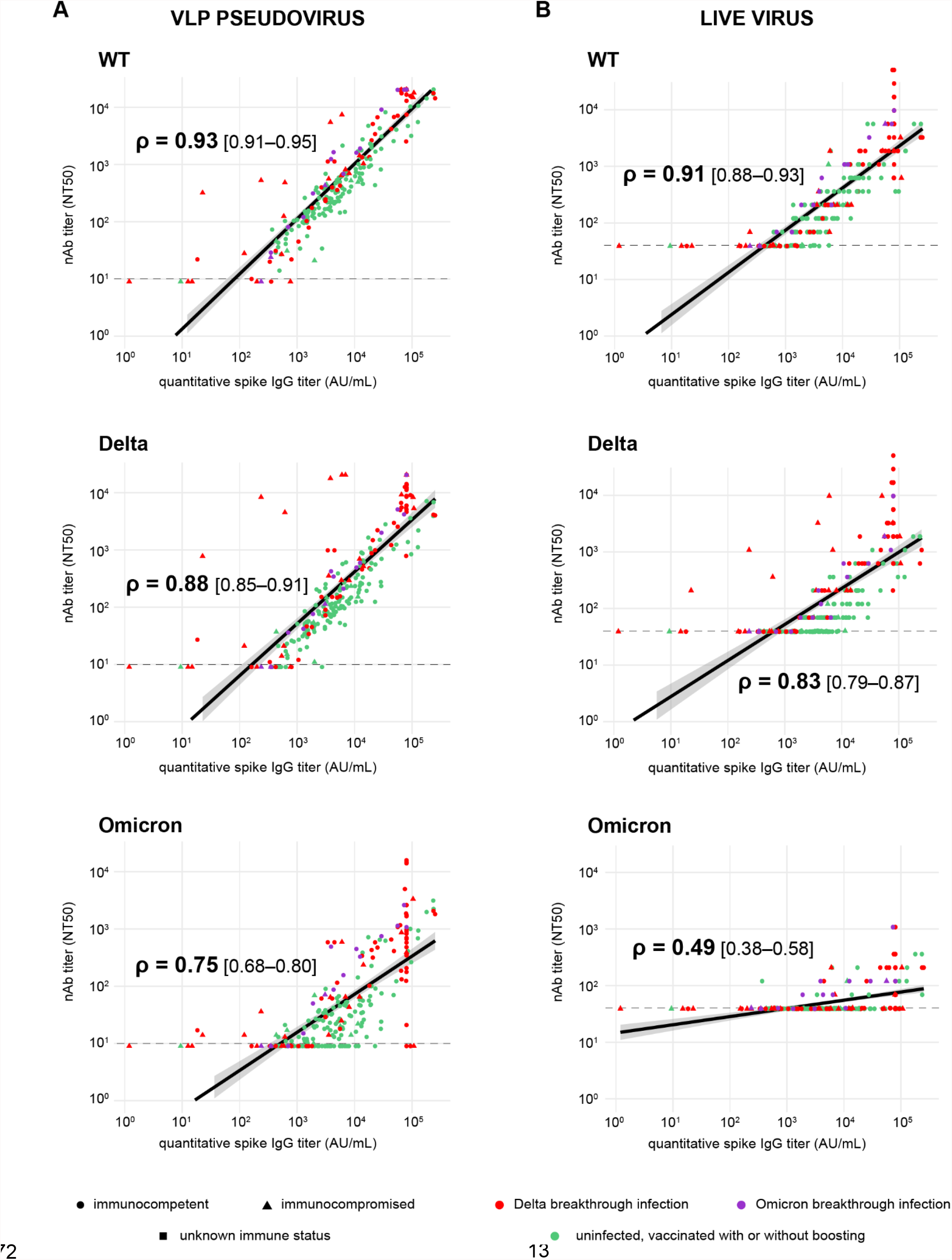
Correlation between quantitative spike IgG and neutralizing antibody titers. **(A)** Plots showing correlation between spike IgG titers and neutralizing antibodies directed against WT (top), Delta (middle) and Omicron (bottom) lineages using a VLP-based assay. **(B)** Plots showing correlation between spike IgG titers and neutralizing antibodies directed against WT (top), Delta (middle) and Omicron (bottom) lineages using a live virus-based assay. The Spearman’s rank coefficient (ρ) was used to assess the strength of correlation.

## Discussion

Here we used VLP and live virus neutralization assays to investigate neutralizing antibody responses in 125 vaccinated individuals, both non-boosted and boosted, and after Delta and Omicron vaccine breakthrough infections. Our results suggest that vaccine boosting and breakthrough infections can restore broad neutralizing hybrid immunity by increasing baseline titers, with higher relative titers against the infecting variant. Notably, Delta-specific titers in Delta breakthroughs rose to become comparable to levels against WT, while Omicron-specific titers in Omicron breakthroughs rose to become comparable to levels against Delta. However, we also found that the magnitude of increase in baseline titers is dependent on the clinical severity of the breakthrough infection. Increased neutralizing antibody levels in Delta related to Omicron breakthrough cases (p=0.049) was likely due to an increased proportion of moderate to severe infections in the Delta compared to the Omicron cohort (p=0.014, 61.5% versus 21.4%). Indeed, moderate to severe clinical infections in both Delta and Omicron breakthrough cohorts were associated with significantly higher neutralization titers than mild or asymptomatic infections among immunocompetent patients (p=0.022).

A limitation of the current study is the lack of both acute and convalescent samples from patients with Delta or Omicron breakthrough infections. Indeed, we found a significant difference in the median days between symptom onset or PCR positivity and sample collection **(Table 1, 17 versus 8**.**5 days, p=0**.**008)**. However, when we analyzed a more comparable subset of samples collected ≤15 days following PCR positivity and sample collection, the difference in neutralization titers between Omicron and Delta was still significant (p=0.037). To confirm these findings, collection and analysis of samples from patients with Omicron breakthrough infections at later time points is ongoing. Other studies have looked at the effect of boosting on neutralization of Omicron and the role cross-variant immunity plays in Omicron breakthrough infections. A study from Laurie, et al. (2022) reported a 4 to 8-fold reduction in neutralization titer in sera from boosted individuals using a pseudovirus assay, comparable to the 7.4-fold reduction that we observed using a VLP assay. Similar to our findings, the study by Khan, et al. (2021) found that sera from patients with Omicron breakthrough infections can enhance Delta virus neutralization to a limited extent (4.4-fold), but that immunity elicited against the specific infecting variant (Omicron) is higher (17.4-fold).

Our findings have implications with regard to the likelihood of Omicron infections providing mass immunization on the population level against SARS-CoV-2. Widespread infections from Omicron globally both in vaccinated and unvaccinated persons, have been reported, and Omicron has shown to cause milder disease with reduced risk of hospitalization and death relative to prior lineages (Wolter *et al*., 2022). In addition, epidemiologic data to date suggests that Omicron may be rapidly outcompeting more pathogenic variants such as Delta (Outbreak.info, 2022). These observations raise the prospect of Omicron being a harbinger of the end of the pandemic as SARS-CoV-2 becomes an endemic virus and broad swaths of the population acquire vaccine-mediated and/or natural immunity. However, we found a significantly smaller rise of neutralization titers associated with milder Omicron breakthrough infection in vaccinated individuals, to only approximately one-third of the rise associated with boosting. We also identified limited cross-variant immunity to Delta. Thus, breakthrough infection from Omicron may enhance cross-protection against Delta, and vice-versa, inasmuch as there is a sufficiently large increase in baseline neutralizing immunity, which appears to be related to the clinical severity of the infection. Our findings parallel those from another study from our group that demonstrated limited cross-variant immunity after milder Omicron variant infection in unvaccinated individuals in a mouse model and in human patients (Suryawanshi *et al*., 2022). Taken together, our results suggest that Omicron-induced immunity may not be sufficient to prevent infection from another, more pathogenic variant, should it emerge in the future. They also highlight the continued importance of vaccine boosters in enhancing immunity, as breakthrough infection alone may not be reliable in eliciting protective titers against re-infection or future infection from different variants. Furthermore, the relative increase in immunity against the infecting variant in breakthrough infections indicates that the use of variant-specific immunogens in vaccine development remains a viable strategy for addressing VOCs that continue to circulate in the population.

Results from the live virus neutralization studies consistently showed lower titers than those using VLPs, which are similar to spike-pseudotyped viruses. The majority of SARS-CoV-2 neutralization studies reported to date have used pseudoviruses because the protocols for running these assays have been reliable, safe, and convenient. Of note, the VLPs used in this study incorporate all of the Omicron-specific mutations found in the structural spike, nucleocapsid, matrix, and fusion proteins (Syed *et al*. 2022), and not only in the spike protein, as is the case for most pseudovirus assays. One possibility for the discrepant neutralization results may be the use of different cell lines for the VLP (293T) and live virus (Vero) assays, although both cell lines are highly susceptible and permissive to SARS-CoV-2 given stable expression of the ACE2 and TMPRSS2 receptors (Hoffmann *et al*. 2020; Case *et al*. 2020). A more likely explanation is that pseudoviruses and VLPs typically only measure the capacity of the virus to enter cells during a single round of infection, whereas live virus assays measure virus infection over several rounds of infection since the reporting endpoints rely on the appearance of cytopathic effect, during which the viruses have already spread from cell-to-cell. Therefore, the reported extent of immune evasion associated with Omicron infection may be underestimated with the use of pseudovirus assays alone. In this study, for example, we found 7.4X decreases in median neutralization titers (4,727⟶635) against Omicron in boosted individuals using the VLP assay but a 21.4X decrease (1,475⟶69) in titers using the live virus assay, corresponding to 93% and 62% of boosted individuals, respectively, having neutralizing antibody levels above an NT50 cutoff of 40.

The utility of FDA authorized serologic assay results as correlates of immune protection with respect to infection from different variants is still under investigation (Gilbert *et al*. 2021). Here we found that spike IgG quantitative and neutralizing antibody results are less correlated with Delta and Omicron infections and thus less predictive of neutralizing immunity. As expected, the degree of correlation was inversely related to the extent of neutralizing antibody evasion associated with the variant. Despite the presence of multiple spike mutations, measured antibody levels of 10^3^–10^4^ for Delta and >10^5^ for Omicron still reliably predicted neutralization. Nevertheless, serologic assays tailored to individual variants or assays directly measuring neutralization will likely be needed for more accurate assessments of neutralizing immunity.

## Data Availability

All data produced in the present study are available upon reasonable request to the authors.

## Acknowledgments

We thank the staff at UCSF Clinical Laboratories and the UCSF Clinical Microbiology Laboratories for help in identifying and aliquoting nasal swab and plasma samples. This work was funded by US CDC Epidemiology and Laboratory Capacity (ELC) for Infectious Diseases Grant 6NU50CK000539 to the California Department of Public Health (COVIDnet) (M-K.M., C.H., D.A.W., C.Y.C.), the Innovative Genomics Institute (IGI) at UC Berkeley and UC San Francisco (J.D., M.O., C.Y.C.), US Centers for Disease Control and Prevention contract 75D30121C10991 (C.Y.C.), the Roddenberry Foundation (M.O.), National Institutes of Health (NIH) grants R37AI083139 (M.O.), R21AI59666 (J.A.D.), and U54HL147127 (M.M.K.), the Howard Hughes Medical Institute (J.A.D.), the Gladstone Institutes (J.A.D. and M.O.), Abbott Laboratories (C.Y.C.), and the Sandler Program for Breakthrough Biomedical Research (C.Y.C.). The funders had no role in study design, data collection and analysis, decision to publish, or preparation of the manuscript. The findings and conclusions in this article are those of the author(s) and do not necessarily represent the views or opinions of the California Department of Public Health or the California Health and Human Services Agency.

## Author contributions

C.Y.C., M.O., J. D., and C.H. conceived and designed the study. C.Y.C, V.S., N.B., and P.S. coordinated the sequencing efforts and laboratory studies. A.S., M.K.M., A.S-G., N.B., V.S., M.G.K., B.S., M.M.K, A.C., P.Y.C, Y.Z., M.R., and J.P. performed experiments. C.Y.C., V.S., N.B., P.S, A.S., M.K.M, A.S-G., J.N., A.G., M.R., J.P., J.H.Jr., C.H. analyzed data. C.Y.C. and V.S. performed genome assembly. V.S., N.B., P.S., J.N., and A.G. collected samples. C.Y.C., V.S., N.B., and P.S. wrote the manuscript. C.Y.C. and V.S. prepared the figures. C.Y.C., V.S., A.S., M.K.M., N.B., P.S., M.G-K., Y.Z., J.N., A.G., J.H.Jr., C.H., and D.A.W. edited the manuscript. C.Y.C. and V.S. revised the manuscript. All authors read the manuscript and agree to its contents.

## STAR Methods

### RESOURCE AVAILABILITY

#### Lead contact

Further information and requests for resources and reagents should be directed to and will be fulfilled by the Lead Contact, Charles Chiu.

#### Materials availability

Passaged aliquots of the cultured SARS-CoV-2 Omicron virus, synthetic VLPs (virus-like particles), and available remaining clinical nasal swab and serum samples are available upon request.

#### Data and code availability

Assembled SARS-CoV-2 genomes in this study were uploaded to GISAID (Shu and McCauley, 2017) (accession numbers pending). Scripting code used for the data analysis and visualization, a table showing deidentified clinical and demographic metadata, and consensus genome FASTA files are available in a Zenodo data repository pending)

#### Human Sample Collection and Ethics Statement

Blood samples were collected through two methods. First, remnant whole blood and plasma samples from patients hospitalized with COVID-19 at UCSF were retrieved from UCSF Clinical Laboratories daily based on availability. Remnant samples were biobanked and retrospective medical chart reviews for relevant demographic and clinical metadata were performed under a waiver of consent and according to protocols approved by the UCSF Institutional Review Board (protocol numbers 10-01116 and 11-05519). Samples were obtained from pediatric and adult patients of all genders. No analyses based on sex or age were conducted.

Second, plasma samples were also collected through the UMPIRE (UCSF EMPloyee and community member Immune REsponse) study, a longitudinal COVID-19 research study focused on collection of prospective whole blood and plasma samples from enrolled subjects to evaluate the immune response to vaccination, with and without boosting, and/or vaccine breakthrough infection. Informed consent for participation in the UMPIRE study and collection of data and samples were obtained according to a protocol approved by the UCSF Institutional Review Board (protocol number 20-33083). The UMPIRE study cohorts included (1) fully vaccinated individuals with either 2 doses of Emergency Use Authorization (EUA) authorized mRNA vaccine (Pfizer or Moderna) or 1 dose of the EUA authorized Johnson and Johnson vaccine. Consented participants came to a UCSF CTSI Clinical Research Service (CRS) Laboratory where their blood was drawn by nurses and phlebotomists. At each visit, two to four 3mL EDTA tubes of whole blood were drawn, and one or two EDTA tubes were processed to plasma from each timepoint. Relevant demographic and clinical metadata from UMPIRE participants were obtained through participant Qualtric surveys performed at enrollment and at each blood draw. Serum samples were heat inactivated at 56°C for 30 mins prior to use in VLP and live virus assays.

#### Clinical Chart Review

The criteria for an infection of moderate severity included hospitalization for COVID-19 pneumonia with an oxygen requirement of >2L of oxygen by nasal cannula or another infectious complication of the disease (e.g. acute renal injury, diarrhea with electrolyte disturbances, necrosis of the extremities, encephalopathy, etc.). The criteria for a severe infection included COVID-19 pneumonia with severe hypoxemia with an oxygen requirement of >6L, including the need for CPAP (continuous positive airway pressure), BIPAP (bilevel positive airway pressure), or intubation with mechanical ventilation, COVID-19 associated end-organ failure, and/or death. Outpatients and hospitalized patients not meeting criteria for moderate to severe infection were classified as having a mild or asymptomatic infection.

#### Viral Whole-Genome Sequencing

Remnant clinical nasopharyngeal/oropharyngeal (NP/OP) swab samples collected in universal transport media or viral transport media (UTM/VTM) were diluted with DNA/RNA shield (Zymo Research, # R1100-250) in a 1:1 ratio (100 μl primary sample + 100 μl shield). The Omega BioTek MagBind Viral DNA/RNA Kit (Omega Biotek, # M6246-03) and the KingFisherTM Flex Purification System with a 96 deep-well head (ThermoFisher, 5400630) were then used for viral RNA extraction. Extracted RNA was reverse transcribed to complementary DNA and tiling multiplexed amplicon PCR was performed using SARS-CoV-2 primers version 3 according to a published protocol (Quick *et al*. 2017). Adapter ligation was performed using the NEBNext^®^ ARTIC SARS-CoV-2 FS Library Prep Kit (Illumina^®^)(New England Biolabs, # E7658L). Libraries were barcoded using NEBNext Multiplex Oligos for Illumina (96 unique dual-index primer pairs) (New England Biolabs, # E6440L) and purified with AMPure XP (Beckman-Coulter, #. Amplicon libraries were then sequenced on either Illumina Miseq or NextSeq 550 as 2×150 paired-end reads (300 cycles).

#### Genome Assembly and Variant Identification

Raw sequencing data were simultaneously demultiplexed and converted to FASTQ files and screened for SARS-CoV-2 sequences using BLASTn (BLAST+ package 2.9.0). Reads containing adapters, the ARTIC and/or VarSkip primer sequences, and low-quality reads were filtered using BBDuk (version 38.87) and then mapped to the Wuhan-Hu-1 SARS-CoV-2 reference genome (National Center for Biotechnology Information (NCBI) GenBank accession number NC_045512.2) using BBMap (version 38.87). Consensus sequences were generated using iVar (version 1.3.1) (Grubaugh *et al*. 2019) and lineages were assigned using Pangolin (Rambaut *et al*. 2020) (version 3.1.17).

#### Serologic testing

SARS-CoV-2-specific antibodies were determined using the Abbott ARCHITECT SARS-CoV-2 IgG (N-based), AdviseDx SARS-CoV-2 IgM (spike receptor-binding domain (RBD)-based), and AdviseDx SARS-CoV-2 IgG II (spike RBD-based) tests according to the manufacturer’s specifications.

#### VLP neutralization assay

For a 6-well, plasmids CoV2-N (0.67), CoV2-M-IRES-E (0.33), CoV-2-Spike (0.0016) and LucT20 (1.0) at indicated mass ratios for a total of 4 µg of DNA were diluted in 200 µL optimem. 12 µg PEI was diluted in 200 µL Opti-MEM and added to plasmid dilution quickly to complex the DNA. Transfection mixture was incubated for 20 minutes at room temperature and then added dropwise to 293T cells in 2 mL of DMEM containing fetal bovine serum and penicillin/streptomycin. Media was changed after 24 hours of transfection and At 48 hours posttransfection, VLP containing supernatant was collected and filtered using a 0.45 µm syringe filter. For other culture sizes, the mass of DNA used was 1 µg for 24-well, 4 µg for 6-well, 20 µg for 10-cm plate and 60 µg for 15-cm plate. Optimem volumes were 100 µL, 400 µL, 1 mL and 3 mL respectively and PEI was always used at 3:1 mass ratio.

Each heat inactivated serum sample was serially diluted from 1:20 to 1:20480 dilution in complete DMEM media prior to incubation (1hr at 37°C) with 40μL VLPs with total volume of 50μL, then plated onto receiver cells (50000 293T ACE2-TMPRSS2 cells). Next day, supernatant was removed and cells were lysed in 20 µL passive lysis buffer (Promega) for 15 minutes at room temperature with gentle rocking. Lysates were transferred to an opaque white 96-well plate and 30 µL of reconstituted luciferase assay buffer was added and mixed with each lysate. Luminescence was measured immediately after mixing using a TECAN plate reader. Neutralization titer (NT50) was estimated by fitting the interpolating the dilution of serum at which 50% infectivity was observed.

#### SARS CoV-2 isolation in cell cultures

Vero E6-TMPRSS2-T2A-ACE2 and Vero-81 were cultured with MEM supplemented with 1x penicillin-streptomycin (Gibco), glutamine (Gibco) and 10% Fetal calf serum (Hyclone). The Vero E6-TMPRSS2-T2A-ACE2 were also supplemented with 10ug/mL puromycin.

The Omicron and Delta variants were isolated from a patient NP swab. To isolate Delta, 200ul of a nasopharyngeal (NP) sample that was previously sequence identified as Delta, was diluted 1:3 in PBS supplemented with 0.75% bovine serum albumin (BSA-PBS) and added to confluent Vero-81 cells in a T25 flask, allowed to adsorb for 1 hour, then additional media was added and the flask was incubated at 37°C with 5% CO2 with daily monitoring for CPE. When 50% CPE was detected, the contents were collected, clarified by centrifugation and stored at -80C as passage 0 stock. Passaged stock of Delta was made by inoculation of Vero-81 confluent T150 flasks with 1:10 diluted p0 stock, monitored and harvested at approximately 50% CPE. Omicron viral stock was similarly produced from a sequence confirmed NP sample using Vero E6-TMPRSS2-T2A-ACE2 in a T25 and harvested at 90% CPE with no subsequent passaging. Both viral stocks were sequenced to confirm lineage and TCID50 was determined by titration.

#### Live virus neutralization assay

CPE endpoint neutralization assays were done following the limiting dilution model using p0 stock of Omicron and p1 stock of Delta in Vero E6-TMPRSS2-T2A-ACE2. Patient plasma was diluted 1:10 in BSA-PBS and heat inactivated at 56C for 30 minutes. Serial 3-fold dilution of plasma were made in BSA-PBS. Plasma dilutions were mixed with 100 TCID50 of each virus diluted in BSA-PBS at a 1:1 ratio and incubated for 1 hour at 37C. Final plasma dilutions in plasma-virus mixture ranged from 1:40 to 1:84480. 100ul of the plasma-virus mixtures were added in duplicate to flat bottom 96-well plates pre-seeded with Vero E6-TMPRSS2-T2A-ACE2 at a density of 2.5 × 104/well and incubated in a 37°C incubator with 5% CO2 until consistent CPE was seen in the virus control (no neutralizing plasma added) wells. Positive and negative controls were included as well as cell control wells and a viral back titration to verify TCID50 viral input. Individual wells were scored for CPE as having a binary outcome of ‘infection” or ‘no infection’ and the IC50 was calculated using the Spearman-Karber method. All steps were done in a Biosafety Level 3 lab using approved protocols.

### Statistical Analyses and Data Visualization

Statistical analyses and data visualization were performed using R (version 4.0.3) and Python (version 3.7.10). Fisher’s exact test was used to evaluate associations of demographic and clinical variables with variant-specific breakthrough infections. Fold decreases in neutralizing activity were measured by comparing median neutralizing antibody titers. Significance testing was performed using the Wilcoxon signed-rank test and Mann-Whitney U test for paired and unpaired samples, respectively. Correlation coefficients were calculated using Spearman’s rank analysis. Plots were generated using ggplot2 (version 3.3.5) in R and seaborn package (version 0.11.0) in Python. All statistical tests were conducted as two-sided at the 0.05 significance level.

## Supplemental Figures

**Figure S1.**
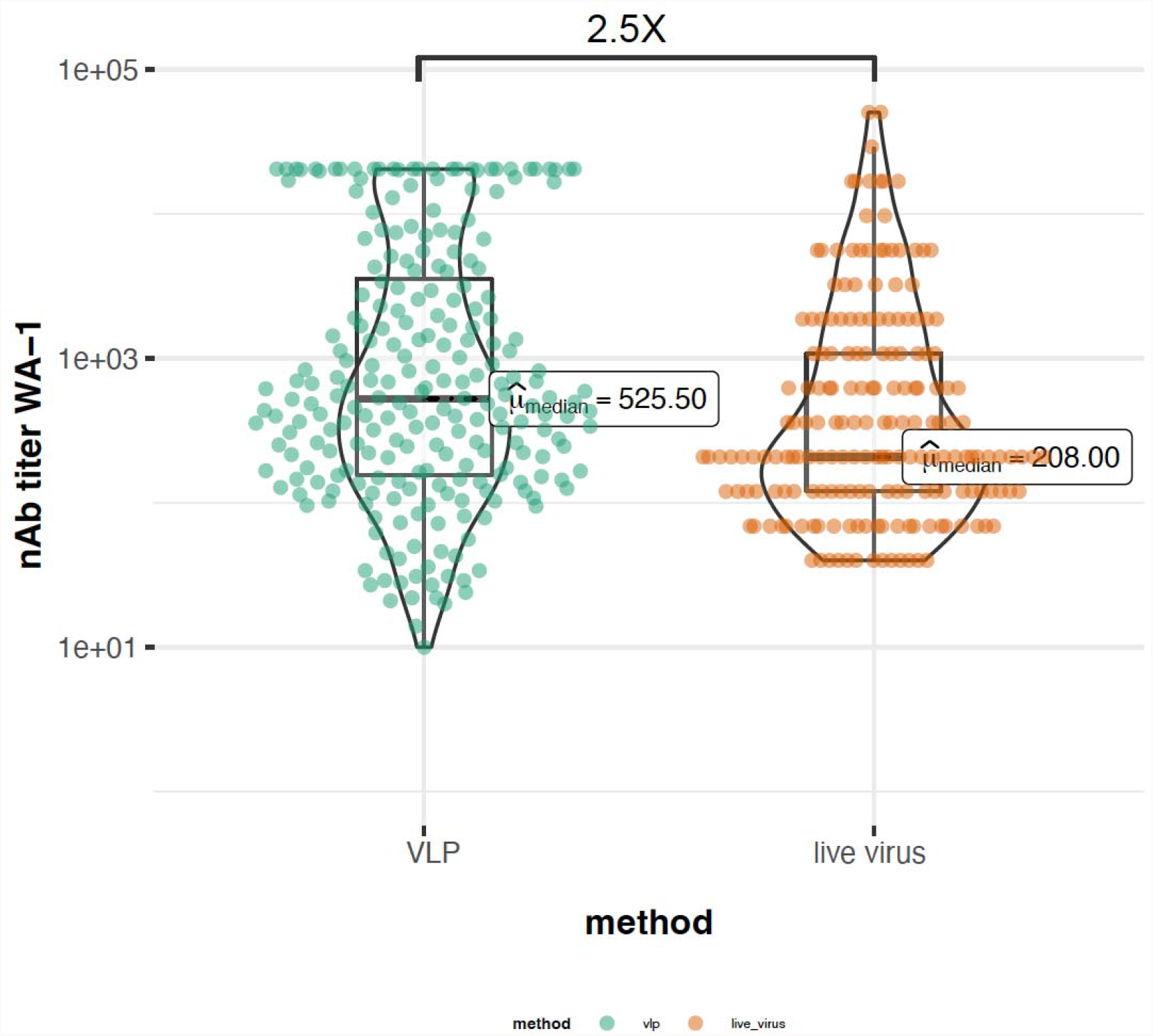
VLP and live virus neutralization assay median neutralizing antibody titers. Plot showing the difference in median neutralizing antibody titers to WT lineage between VLP-based and live virus-based assay.

